# Association of left atrial structure and function with cognitive function among adults with metabolic syndrome

**DOI:** 10.1101/2022.03.11.22272280

**Authors:** Ines Gonzalez Casanova, Ángel M. Alonso-Gómez, Dora Romaguera, Estefanía Toledo, Elena Fortuny, Luis López, Raúl Ramallal, Jordi Salas-Salvadó, Lucas Tojal-Sierra, Olga Castañer, Alvaro Alonso

**Author notes:** Corresponding author: Dr. Inés González-Casanova. Department of Applied Health Science, Indiana University Bloomington School of Public Health, 1065 7^th^ Street, Suite 166, Bloomington, IN, USA 47404.

## Abstract

**Background:** Atrial fibrillation has been associated with cognitive impairment. Whether subclinical abnormalities in atrial function and substrate predisposing to atrial fibrillation impact cognitive function has received limited attention.

**Methods:** We tested associations of echocardiographic markers of atrial structure and function with cognitive functioning and decline among 510 participants with overweight/obesity and metabolic syndrome (mean age [standard deviation] of 64.4 [5.2] years in men and 66.5 [3.9] in women). Left atrial markers (volume index, emptying fraction, strain, function index, and stiffness index) were estimated based on transthoracic echocardiography. General cognitive functioning (Mini-mental examination), verbal ability (verbal fluency test), memory and attention (Digit Span Tests), and processing speed and executive function (Trail-Making Tests A and B) were assessed at baseline and at the two-year follow-up. Multiple linear regression was used to test associations of atrial markers (modeled in standard deviation units) with baseline and two-year changes in cognitive scores adjusting for demographic and health covariates.

**Results:** Left atrial structure and function was not associated with cognitive function at baseline. Larger left atrial volume index (standardized β [95% confidence interval] = -0.13 [-0.22, -0.03]), lower peak longitudinal strain (−0.11 [-0.20, -0.01]), and higher stiffness index (−0.18 [-0.28, -0.08) were associated with 2-year worsening in Trail-Making Test A. Strain measurements were also associated with 2-year change in the Controlled Oral Word Association Test.

**Conclusion:** Overall, adverse markers of left atrial structure and function were associated with 2-year detrimental executive function-related cognitive changes in a sample of participants at high risk for cardiovascular disease, highlighting left atrial substrate as a potential risk factor for cognitive decline and dementia.

## Introduction

Atrial fibrillation (AF), a common cardiac arrhythmia associated with high risk of cardiovascular events, has been identified as an important risk factor for age-related mild cognitive decline and dementia.^1,2^ In 2010, AF affected 6.1 million people in the US and it is expected to rise to 12.1 million in 2030; it has been estimated that approximately 1 in 3 people will develop atrial fibrillation in their lifetime. Similarly, 6 to 8% of US adults over 60 years suffer from dementia and 37% have age-related mild cognitive impairment.^3^ While a breadth of evidence supports AF as an important risk factor for cognitive impairment and dementia,^2,4^ the specific pathways linking these diseases are not yet well-understood.

The increased risk of cognitive impairment among people with AF is independent of shared risk factors.^5^ Specific causal pathways potentially responsible for this association include increased risk of cerebral infarcts, microhemorrhages and cerebral hypoperfusion in people with AF, as well as a general pro-inflammatory state in AF, although this latter mechanism remains poorly understood.^5^ Also, the increased risk of cognitive impairment in AF might be a consequence of abnormalities in the atrial structural and functional substrate, independently of the presence of the arrhythmia.^6^

The pathophysiology of AF includes an electrical component that acts on a vulnerable structural and functional atrial substrate, which facilitates the onset and perpetuation of this arrythmia.^7^ In this context, markers of atrial remodeling and fibrosis, such as enlarged left atrial (LA) size and volume, and abnormal LA function, are predictors of AF onset.^8^ Prior studies suggest that LA abnormalities identified through the electrocardiogram are associated with an increased risk of dementia independently of AF.^9^ Similarly, LA volume has been associated with cognitive decline in older adults.^10^ The association of echocardiographic markers of LA function with cognitive function and dementia, however, has received limited attention. Therefore, the objective of this analysis was to assess whether markers of LA structure and function, which provide the AF substrate, were associated with cognitive function and cognitive decline among overweight or obese adults with metabolic syndrome participating in the PREDIMED-PLUS randomized trial.

## Methods

### Study population

The PREDIMED-PLUS trial is an ongoing multicenter randomized controlled trial aimed at preventing cardiovascular disease in overweight or obese adults with metabolic syndrome (ISRCTN89898870). Recruitment started in 2013 and included 3,574 men (age 55-75 y) and 3,300 women (age 60-75 y). Participants were recruited in 23 centers in Spain and randomized 1:1 to an intensive lifestyle intervention (ILI) program based on an energy-restricted Mediterranean diet, increased physical activity, and cognitive-behavioral weight management or to a control intervention of low-intensity dietary advice on the Mediterranean diet (not energy-restricted).^11^

Inclusion criteria for the PREDIMED-PLUS trial were men aged 55–75 years and women aged 60–75 years, with overweight or obesity (body mass index 27–40 kg/m^2^), who at baseline met at least three components of the metabolic syndrome. Exclusion criteria have been described elsewhere and included documented history of cardiovascular disease among others.^12^ For this analysis, we included a sub-sample of 510 PREDIMED-PLUS participants from three recruiting centers (University of Navarra, Araba University Hospital, Son Espases University Hospital) who underwent transthoracic echocardiography at baseline and who also had information on at least one cognitive test at baseline or at the two-year follow-up.

Study protocols were approved by Institutional Review Boards at all participating institutions. All participants provided written informed consent.

### Cognitive function assessments

At baseline and at the two-year follow-up visit, participants underwent a detailed cognitive assessment evaluating different domains.

The Mini-mental state examination (MMSE)^13,14^ test was validated in Spanish population to assess general cognitive impairment. It is a 30-point questionnaire that produces sub-scales for time and spatial orientation, immediate and deferred recall, attention, calculation, and language. These scores are added to produce the overall MMSE score, where a threshold of less than 24 is defined as cognitive impairment.^15^

We used the Spanish version of the Digit Span Test^16^ that is part of the Wechsler Adult Intelligence Scale battery.^17^ It has a forward component that measures short-term memory and attention, and a backward component that measures working memory. In the forward test, participants are requested to repeat a series of random single digits in the order they heard them, and in the backward test, they are requested to repeat a series of numbers in the inverse of the order they heard them.

Semantic and phonetic fluency were respectively evaluated by asking participants to mention as many animals or words containing the letter “P” as possible in 60 seconds.^18,19^

Finally, the Trail Making Test^20^ measures processing speed and executive function. The Trail Making Test A, where participants are asked to connect numbers 1 to 25 in the correct order, is meant to assess cognitive processing skills. The task B consists of connecting numbers and letters alternatively in the correct order.

### Echocardiographic assessment

Participants from three sites were invited to undergo transthoracic echocardiography following a common protocol. Echocardiographic examinations were performed at baseline using an ultrasound scanner Vivid 7 or Vivid 9 (General Electric Healthcare). M-mode, doppler imaging and two-dimensional cine loops for 3 heart beats of standards views were obtained from each patient. All images were then digitally stored. The offline ultrasound software EchoPac (GE Healthcare) was used for image analysis and measurements. For reliability purposes, images were read at a core reading center (Son Espases University Hospital) by two trained readers blind to clinical data following recommendations of the American Society of Echocardiography.^21^ Markers of LA structure included the LA volume index (LA volume indexed to body surface area), while markers of LA function included the LA emptying fraction, the peak LA longitudinal strain, conduit strain, and contractile strain, the LA function index, and the LA stiffness index. LA emptying fraction was calculated as *(V*_*max*_*-V*_*min*_*)/V*_*max*_, where V_max_ is the maximum volume of the LA just before the opening of the mitral valve and V_min_ is the minimal volume at the closure of the mitral valve. LA emptying fraction is a measure of the LA volumetric function. The peak LA longitudinal strain, a marker of LA myocardial function, was measured at the end of the reservoir phase. The conduit and contractile strains relate to LA function during the diastole: early diastole strain represents conduit function and late diastole strain represents contractile function. The LA function index combines the reservoir function, the adjusted LA volume, and the stroke volume to reflect left ventricular systolic and diastolic function. It was calculated as (LA emptying fraction × left ventricular outflow tract-velocity time integral) / indexed LA end-systolic volume.^22^ The LA stiffness index, a measure of LA remodeling and fibrosis, was calculated as E/e’ ratio divided by peak LA systolic longitudinal strain, where E represents the early mitral inflow velocity (E wave) and e’ represents the medial and lateral mitral annular velocity.^23^

### Other Covariates

Participants’ sociodemographic and health characteristics were assessed at baseline via a face-to-face questionnaire. Sociodemographic variables included age at baseline, sex, marital status (single or married), education (years of schooling), occupation (employed, unemployed, retired, housework). Body mass index was measured at baseline and included in the analysis, as well as self-reported history of diabetes, hypertension, depression, or non-AF arrythmia. CHA2DS2-VASc score was calculated using baseline information and was used as a descriptive variable.^24^ The recruitment site (Mallorca, Vitoria, or Navarra) was also included as a covariate in this analysis.

### Statistical Analysis

We first conducted exploratory analysis by testing bi-variate unadjusted associations between markers of LA structure and function and cognitive scores at baseline, as well as associations with baseline sociodemographic variables using Spearman or Pearson correlations, t-tests and ANOVAs as appropriate. LA structure and function variables were scaled in standard deviation units to facilitate comparisons across variables. For LA conduit and contractile strain, we used the absolute value instead of the negative percentage to keep the direction of our estimates consistent with peak LA longitudinal strain estimates. All cognitive functioning scores were standardized to a mean of 0 and a standard deviation of 1 using the mean and standard deviation of the sample. With the exception of the Trail Making Test, lower values of the cognitive tests reflect worse cognitive function. To ensure consistency in the interpretation of results, the analyses used the additive inverse of Trail Making Tests score [-(Trail Making Test)] to have lower values in this test indicate worse test performance.

Multivariable linear models were used to test cross-sectional associations between markers of LA structure and function and cognitive functioning outcomes at baseline. We ran models adjusted only for age, sex and site, and then fully adjusted for sociodemographic characteristics (marital status, years of schooling, employment), medical history (diabetes, hypertension, minor arrythmias, depression), body mass index, and smoking (categorized into current, former, or never smoker), 17-item Mediterranean Diet score,^25^ and METs per day of moderate to vigorous physical activity estimated using the long version of the Minnesota leisure time physical activity questionnaire.^26^ Estimates where the 95% confidence interval did not cross the null in both partially and fully adjusted models were considered significant.

The difference between cognitive function scores at baseline and at the 2-year follow-up was calculated by subtracting the former from the latter. Multivariate linear regression was used to test the association between LA structure and function and this difference using a similar modeling approach that was additional adjusted for intervention arm as a potential confounder.

A sensitivity analyses was performed after excluding participants that were born outside of Spain (n=20) because Spanish may not be their first language. An additional sensitivity analysis excluding those with MMSE <24 had been planned but none of the participants had values below this cutoff.

## Results

There were 510 participants included in the analyses, of which 468 (92%) had a repeated cognitive assessment two years later (Figure 1). Participants were on average 65 years at baseline, and majority male (60%). The mean body mass index was 32 kg/m^2^. Men were significantly more likely to smoke or be former smokers than women and were more physically active. Women had fewer years of schooling and were more likely to have depression (Table 1).

**Table 1.**

| **% or mean (SD)** | **Male (307)** | **Female (203)** | **Total (510)** |
| --- | --- | --- | --- |
| Site |  |  |  |
| Mallorca | 24.1 | 36.5 | 29.0 |
| Navarra | 19.5 | 18.7 | 19.2 |
| Vitoria | 56.3 | 44.8 | 51.8 |
| Age (years) | 64.4 (5.2) | 66.5 (3.9) | 65.1 (4.8) |
| Married | 85.0 | 65.5 | 77.3 |
| People living in the household | 1.4 (1.0) | 1.1 (0.9) | 1.3 (1.0) |
| Schooling (years) | 13.0 (5.5) | 10.4 (4.4) | 12.0 (5.3) |
| Employed | 21.5 | 9.9 | 16.9 |
| Health status |  |  |  |
| Body Mass Index (kg/m<sup>2</sup>) | 32.4 (3.1) | 32.9 (3.6) | 32.6 (3.3) |
| Diabetes | 22.8 | 25.1 | 23.7 |
| Hypertension | 84.4 | 81.8 | 83.2 |
| Depression | 13.0 | 28.1 | 19.0 |
| Arrhythmia | 5.9 | 5.9 | 5.9 |
| CHA<sub>2</sub>DS<sub>2</sub>-VASc Score for Atrial Fibrillation | 1.6 (0.8) | 2.8 (0.9) | 2.1 (1.0) |
| Stroke Risk |  |  |  |
| Health Behavior |  |  |  |
| Current Smoker | 10.8 | 7.2 | 9.4 |
| Former Smoker | 66.8 | 29.1 | 51.8 |
| Mediterranean Diet Score | 7.2 (2.7) | 8.0 (3.0) | 7.5 (2.9) |
| Moderate to vigorous physical activity (METs/day) | 332.9 (360.6) | 173.5 (213.0) | 269.4 (319.8) |
| General Echocardiographic Measures |  |  |  |
| Left Ventricle Mean Ejection Fraction (%) | 64.9 (6.3) | 66.1 (7.9) | 65.4 (7.0) |
| Left Atrial Substrate |  |  |  |
| Volume Index (mL/m<sup>2</sup>) | 23.3 (7.2) | 23.1 (8.2) | 23.2 (7.6) |
| Emptying Fraction (%) | 58.6 (17.8) | 55.9 (10.0) | 57.5 (15.2) |
| Peak Systolic Longitudinal Strain (%) | 27.8 (6.9) | 26.4 (6.6) | 27.2 (6.8) |
| Conduit Strain (%) | -12.2 (4.2) | -11.5 (4.6) | -11.9 (4.4) |
| Contractile Strain (%) | -15.7 (5.3) | -14.8 (4.6) | -15.3 (5.2) |
| Function Index (U) | 64.3 (41.0) | 66.2 (28.8) | 65.1 (36.6) |
| Stiffness Index (U) | 0.3 (0.2) | 0.4 (0.2) | 0.4 (0.2) |

**Figure 1.**
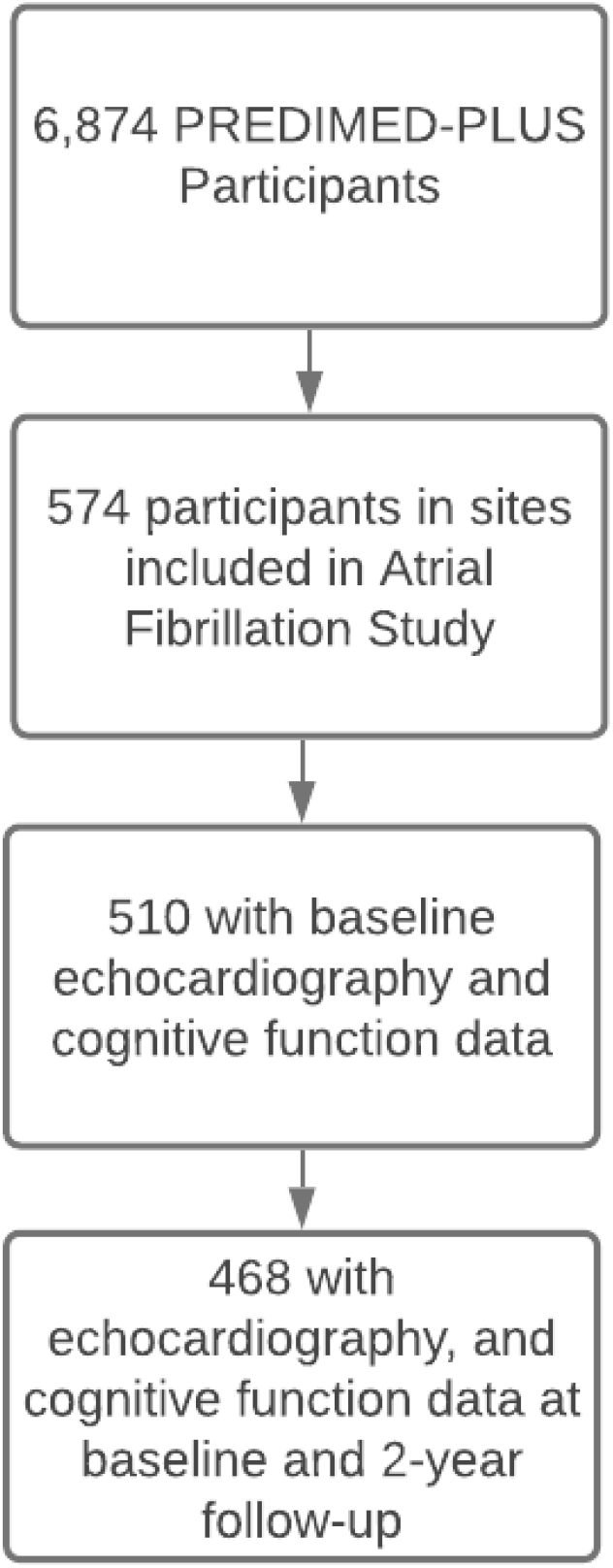
Flowchart of participants included for this analysis.

The mean (±standard deviation) MMSE scores at baseline were 28.9 (±1.5) and 27.9 (±2.2) for men and women, respectively. Among men, there was a small decrease of 0.1 between baseline MMSE score and two-year follow up, while among women there was an increase of 0.1 (Table 2). Similarly, small average decreases were found for semantic fluency, digit span and the trail making tests a and b; these negative differences were greater among men.

**Table 2:**
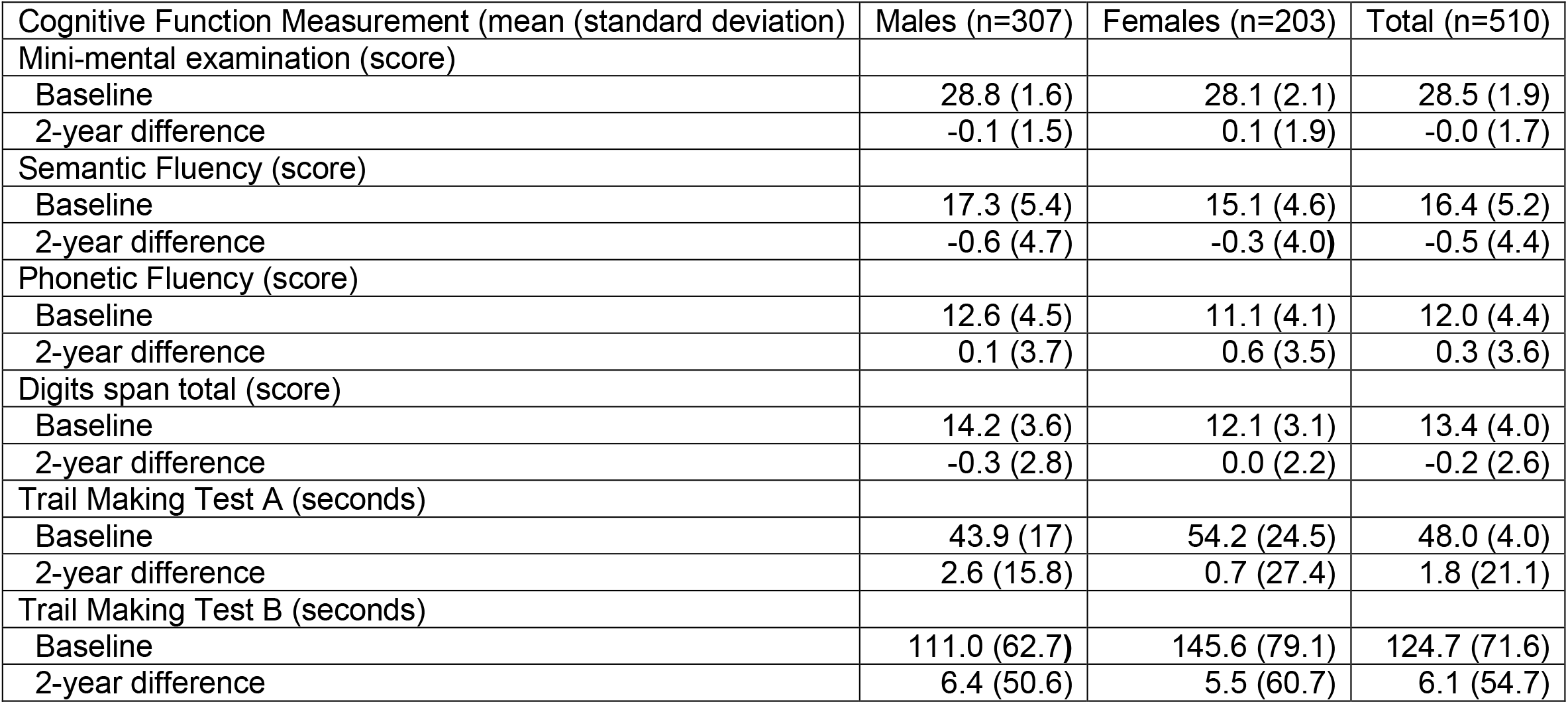
Cognitive function at baseline and 2-year difference among 510 participants of the PREDIMED+ study.

| Cognitive Function Measurement (mean (standard deviation)) | Males (n=307) | Females (n=203) | Total (n=510) |
| --- | --- | --- | --- |
| Mini-mental examination (score) |  |  |  |
| Baseline | 28.8 (1.6) | 28.1 (2.1) | 28.5 (1.9) |
| 2-year difference | -0.1 (1.5) | 0.1 (1.9) | -0.0 (1.7) |
| Semantic Fluency (score) |  |  |  |
| Baseline | 17.3 (5.4) | 15.1 (4.6) | 16.4 (5.2) |
| 2-year difference | -0.6 (4.7) | -0.3 (4.0) | -0.5 (4.4) |
| Phonetic Fluency (score) |  |  |  |
| Baseline | 12.6 (4.5) | 11.1 (4.1) | 12.0 (4.4) |
| 2-year difference | 0.1 (3.7) | 0.6 (3.5) | 0.3 (3.6) |
| Digits span total (score) |  |  |  |
| Baseline | 14.2 (3.6) | 12.1 (3.1) | 13.4 (4.0) |
| 2-year difference | -0.3 (2.8) | 0.0 (2.2) | -0.2 (2.6) |
| Trail Making Test A (seconds) |  |  |  |
| Baseline | 43.9 (17) | 54.2 (24.5) | 48.0 (4.0) |
| 2-year difference | 2.6 (15.8) | 0.7 (27.4) | 1.8 (21.1) |
| Trail Making Test B (seconds) |  |  |  |
| Baseline | 111.0 (62.7) | 145.6 (79.1) | 124.7 (71.6) |
| 2-year difference | 6.4 (50.6) | 5.5 (60.7) | 6.1 (54.7) |

At baseline, none of the LA structural and functional substrate markers were cross-sectionally associated with cognitive functioning tests (Table 3). Among 468 participants with repeated cognitive assessment included in the longitudinal analysis, however, higher LA peak systolic longitudinal strain was associated more favorable 2-year changes in the Digits Span Test and Trail Making Test A. Per each standard deviation increase in longitudinal strain, there was a 0.1 standard deviation increase in the 2-year difference for both tests after adjusting for all sociodemographic and health covariates (Table 4). Larger LA volume index and higher LA stiffness index were both associated with more decline in Trail Making Test A (−0.13, 95% CI -0.22, -0.03 and -0.18, 95%CI -0.28, -0.08, respectively). An inverse association was observed between conduit strain and phonetic fluency, with higher absolute conduit strain associated with larger declines in phonetic fluency (−0.10, 95%CI -0.17, -0.02) (Table 4).

**Table 3:** Association of left atrium structure and function with baseline cognitive function scores among 510 participants of the PREDIMED-PLUS Trial.

| Left atrial measurements |  | Mini-mental examination | Semantic fluency | Phonetic fluency | Digits span total (n=389) | Trail Making Test A | Trail Making Test B |
| --- | --- | --- | --- | --- | --- | --- | --- |
| Volume Index | Model 1 | -0.03<br>(-0.12, 0.06) | 0.06<br>(-0.03, 0.15) | -0.03<br>(-0.13, 0.06) | 0.01<br>(-0.09, 0.10) | 0.07<br>(-0.02, 0.15) | 0.02<br>(-0.06, 0.11) |
|  | Model 2 | -0.03<br>(-0.12, 0.06) | 0.05<br>(-0.04, 0.15) | -0.05<br>(-0.14, 0.04) | 0.01 (-0.09, 0.11) | 0.06<br>(-0.03, 0.15) | 0.02<br>(-0.07, 0.10) |
| Emptying Fraction | Model 1 | -0.04<br>(-0.12, 0.05) | -0.04<br>(-0.12, 0.04) | 0.04<br>(-0.04, 0.12) | -0.01<br>(-0.10, 0.08) | -0.01<br>(-0.09, 0.07) | -0.04<br>(-0.12, 0.05) |
|  | Model 2 | -0.03 (-0.12, 0.05) | -0.05 (-0.14, 0.03) | 0.01 (-0.07, 0.09) | -0.01 (-0.10, 0.08) | -0.01 (-0.10, 0.07) | -0.05 (-0.13, 0.03) |
| Peak Systolic Longitudinal Strain | Model 1 | -0.01<br>(-0.09, 0.08) | -0.04<br>(-0.13, 0.04) | -0.03<br>(0.12, 0.05) | -0.06<br>(-0.16, 0.03) | -0.02<br>(-0.10, 0.06) | -0.03<br>(-0.11, 0.06) |
|  | Model 2 | -0.02 (-0.11, 0.06) | -0.05 (-0.14, 0.03) | -0.03 (-0.12, 0.05) | -0.10 (-0.19, 0.00) | -0.06 (-0.14, 0.03) | -0.06 (-0.14, 0.02) |
| Conduit Strain | Model 1 | -0.03<br>(-0.11, 0.05) | 0.00<br>(-0.08, 0.08) | 0.04<br>(-0.12, 0.13) | 0.01<br>(-0.08, 0.11) | 0.00<br>(-0.08, 0.08) | -0.01<br>(-0.09, 0.07) |
|  | Model 2 | -0.04 (-0.13, 0.03) | -0.01 (-0.09, 0.07) | 0.02 (-0.07, 0.10) | -0.02 (-0.11, 0.07) | -0.02 (-0.10, 0.06) | -0.03 (-0.11, 0.06) |
| Contractile Strain | Model 1 | 0.00<br>(-0.08, 0.08) | -0.06<br>(-0.14, 0.02) | -0.05<br>(-0.13, 0.03) | -0.08<br>(-0.17, 0.01) | -0.03<br>(-0.11, 0.05) | -0.02<br>(-0.09, 0.06) |
|  | Model 2 | -0.01 (-0.09, 0.07) | -0.06 (-0.14, 0.02) | -0.06 (-0.15, 0.02) | -0.11 (-0.20, -0.02) | -0.05 (-0.13, 0.03) | -0.05 (-0.13, 0.03) |
| Function Index | Model 1 | -0.01<br>(-0.10, 0.07) | -0.04<br>(-0.13, 0.04) | 0.00<br>(-0.08, 0.09) | -0.01<br>(-0.10, 0.09) | -0.03<br>(-0.11, 0.06) | -0.05<br>(0.13, 0.04) |
|  | Model 2 | -0.01 (-0.10, 0.08) | -0.04 (-0.12, 0.04) | 0.05 (-0.04, 0.14) | -0.01 (0.10, 0.09) | -0.03 (-0.12, 0.05) | -0.05 (-0.13, 0.03) |
| Stiffness Index | Model 1 | -0.06<br>(-0.14, 0.03) | 0.00<br>(-0.09, 0.08) | -0.01<br>(-0.09, 0.08) | 0.07<br>(-0.03, 0.17) | 0.01<br>(-0.09, 0.07) | -0.01<br>(-0.09, 0.07) |
|  | Model 2 | -0.06 (-0.15,<br>0.03) | -0.01 (-0.09,<br>0.08) | 0.01 (-0.08,<br>0.10) | 0.09 (-0.01,<br>0.19) | 0.07 (-0.01,<br>0.16) | 0.02 (-0.06,<br>0.10) |

Model 1: Adjusted by age, sex and site. Model 2: Adjusted by age, sex, site, number of people living in the household, employment status (yes or no), years of schooling, current smoking (yes or no) and former smoker (yes or no), METS per day of moderate to vigorous physical activity, Mediterranean diet 17-item score, body mass index, diagnosis of arrhythmia, diagnosis of diabetes, and diagnosis of depression at baseline. Values are standardized betta coefficients representing standard deviation change in cognitive function outcomes per a standard deviation increase in left atrial measurements.

**Table 4:**
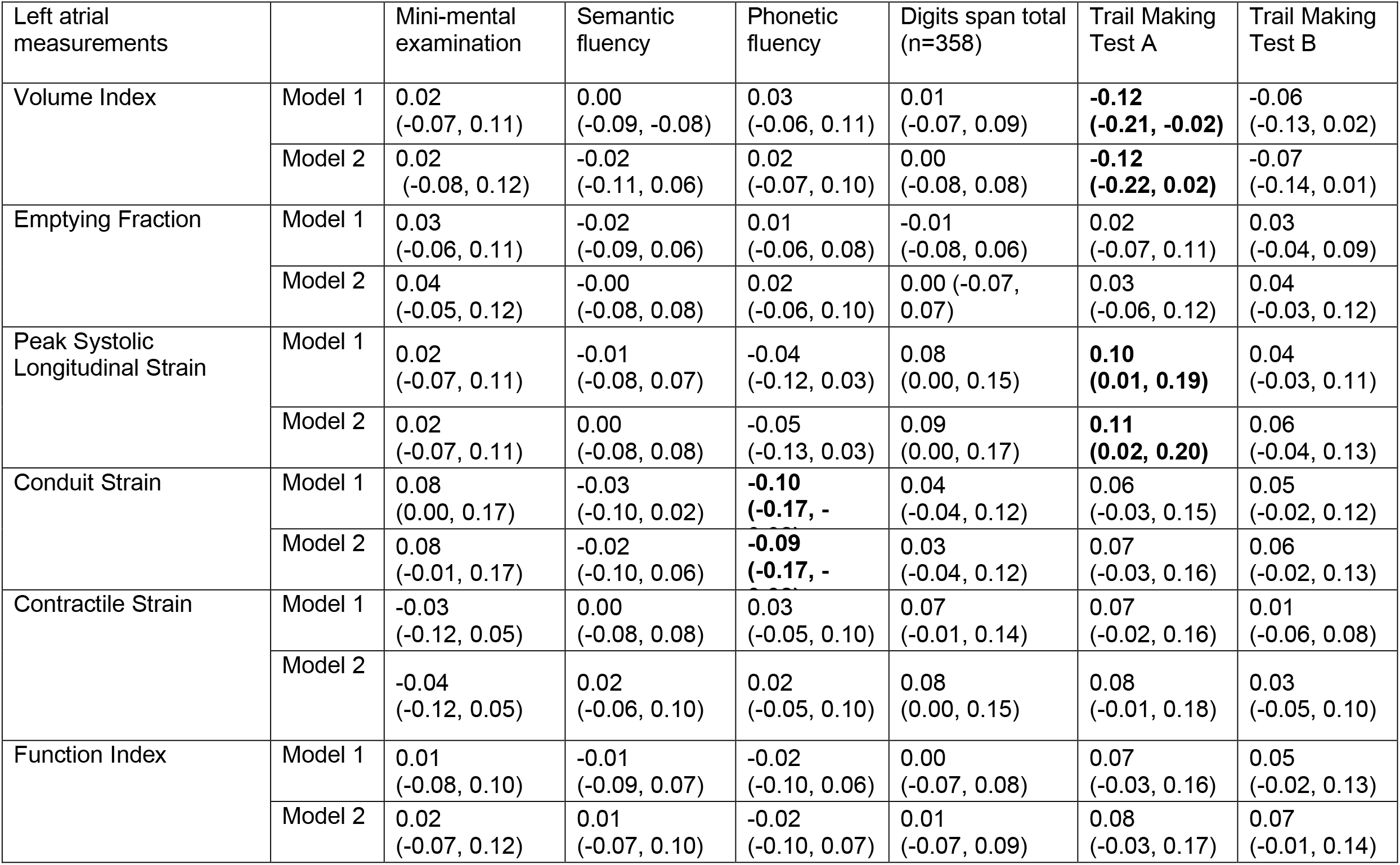
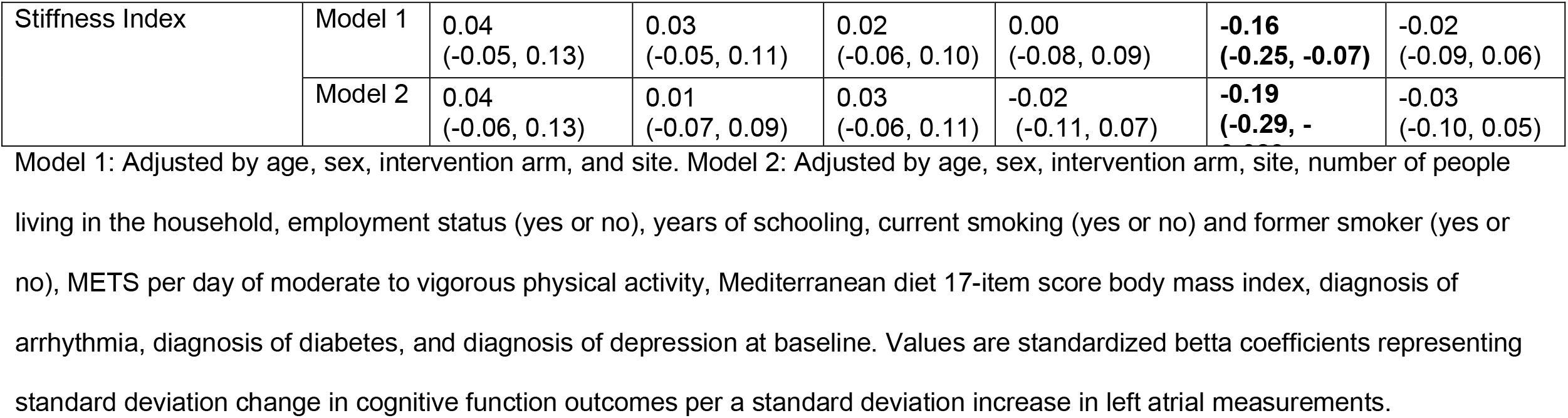
Association of markers of left atrium structure and function with 2-year difference cognitive function scores among 468 participants of the PREDIMED-PLUS Trial.

The sensitivity test excluding foreign born participants did not modify the results.

## Discussion

Previous studies have shown associations between cardiovascular risk in general or AF in particular, and cognitive decline or dementia.^2,4,27^ The aim of this study was to test associations between markers of LA remodeling, which are associated with AF, and cognitive function and change in a sample of overweight adults with no cardiovascular disease or dementia. We found no associations of markers of LA structure and function (volume, emptying fraction, strain, function index, and stiffness index) with various measurements of cognitive function at baseline. We did, however, find some associations between markers of favorable LA volume and function (lower LA volume index, higher LA peak longitudinal strain, lower LA stiffness index) and more favorable 2-year changes in scores of tests related to executive function (Trail Making Test A, Digits Span Test) among this subsample of participants of the PREDIMED-PLUS intervention study.

A large body of evidence supports an association between AF and cognitive decline. In turn, measures of LA structure and function, which characterize the LA substrate of AF, have been associated with increased risk of AF and cardiovascular disease.^28^ Less evidence is available of the potential impact of the AF substrate on cognitive function, although some studies have suggested that even subclinical cardiac dysfunction and markers of AF risk could be associated with cognitive decline in older adults. ^9,29,30^

Left atrial volume index has been widely used as a marker of cardiac structural health, where higher values (LA dilatation) predict worse cardiovascular outcomes. ^31^ Prior work has also evaluated the association of LA volume with cognitive outcomes. A previous study found associations between LA volume index and cognitive impairment defined using a cutoff of <25 in the Mini-mental examination in older adults.^10^ However, a more recent study found that LA enlargement measured by volume index >34 was not associated with 5-year decline in a composite cognitive score.^32^ In our analysis, we found an inverse association between LA volume index and 2-year decline in the Trail Making Test A, but not in the Mini Mental Examination or in other domains. This is a sample of participants with no severe cognitive impairment at baseline (based on the trial exclusion criteria) and with a shorter follow-up. It is possible that the Trail Making Test is more sensitive to small changes related to subclinical alterations in volume and function because it measures executive functioning and has been directly linked to activity in the prefrontal cortex.^33^

Peak atrial longitudinal strain is a measure of LA function, where lower values reflect impaired reservoir function. We found that higher peak LA longitudinal strain was associated with less decline in executive function, specifically in the Trail-Making Test A. Conversely, conduit strain, a subcomponent of the longitudinal strain and a marker of LA conduit function, was inversely associated with phonetic scores. The direction of this finding is inconsistent with the LA longitudinal strain results, making us cautious in the overall interpretation of the results. Left atrial stiffness index is a marker of atrial remodeling and indicates deteriorated reservoir function. It is calculated non-invasively as the quotient of the E/e’ ratio and the LA peak longitudinal strain ^34^. In our study, it was associated with worse decline in the Trail Making Test A, a measure of executive function. To our knowledge this is the first study to find associations between this index and cognitive function, and further research is necessary to confirm this association.

We should also consider multiple testing when interpreting our results. Among many tests conducted, even if most of them were consistent in terms of direction of association and magnitude of effect, only a few reached the traditional threshold of statistical significance, raising concerns that some or all could be chance findings due to multiple testing. Hence, while the results contribute to the general body of evidence in the field, they require confirmation in separate studies.

Strengths of the study include the careful assessment of LA structure and function using echocardiography, the relatively large sample size, the excellent retention of study participants over time, the various measurements of different aspects of cognitive function and repeated measurements that allowed assessing differences over time, and information on a wide range of covariates.

The PREDIMED-PLUS trial excluded people with cardiovascular disease or with severe cognitive functioning from enrollment, which may have limited our ability to study a higher-risk population and may have affected the baseline analyses. Another limitation of our study is that cognitive function was only available at two time points and that the follow-up time between the assessments was only 2 years. Future studies should assess the association between AF substrate and cognition among populations with higher prevalence of AF and wider variation on cognitive function, as well as the association with cognitive decline after longer follow-ups.

In conclusion, better LA structure and function were associated with lower decline in cognitive tests reflecting the areas of memory and executive function in individuals at high risk of cardiovascular disease, highlighting the importance of the LA substrate as a potential risk factor for cognitive decline and dementia.

## Data Availability

Data are not available because the trial is still ongoing.

## References

1. Santangeli P, Di Biase L, Bai R, Mohanty S, Pump A, Cereceda Brantes M, Horton R, Burkhardt JD, Lakkireddy D, Reddy YM, et al. Atrial fibrillation and the risk of incident dementia: A meta-analysis. Heart Rhythm. 2012;9:1761-1768.e1762. doi: http://dx.doi.org/10.1016/j.hrthm.2012.07.026

2. Islam MM, Poly TN, Walther BA, Yang H-C, Wu CC, Lin M-C, Chien S-C, Li Y-C. Association Between Atrial Fibrillation and Dementia: A Meta-Analysis. Frontiers in aging neuroscience. 2019;11:305–305. doi: 10.3389/fnagi.2019.00305

3. Plassman BL, Potter GG. Epidemiology of dementia and mild cognitive impairment. In: APA handbook of dementia. Washington, DC, US: American Psychological Association; 2018:15–39.

4. Kalantarian S, Stern TA, Mansour M, Ruskin JN. Cognitive impairment associated with atrial fibrillation: a meta-analysis. Annals of internal medicine. 2013;158:338–346. doi: 10.7326/0003-4819-158-5-201303050-00007

5. Madhavan M, Graff-Radford J, Piccini JP, Gersh BJ. Cognitive dysfunction in atrial fibrillation. Nature Reviews Cardiology. 2018;15:744–756. doi: 10.1038/s41569-018-0075-z

6. Alonso A, Arenas de Larriva AP. Atrial Fibrillation, Cognitive Decline And Dementia. European cardiology. 2016;11:49–53. doi: 10.15420/ecr.2016:13:2

7. Markides V, Schilling RJ. Atrial fibrillation: classification, pathophysiology, mechanisms and drug treatment. Heart. 2003;89:939. doi: 10.1136/heart.89.8.939

8. Gallinoro E, D’Elia S, Prozzo D, Lioncino M, Natale F, Golino P, Cimmino G. Cognitive Function and Atrial Fibrillation: From the Strength of Relationship to the Dark Side of Prevention. Is There a Contribution from Sinus Rhythm Restoration and Maintenance? Medicina (Kaunas, Lithuania). 2019;55:587. doi: 10.3390/medicina55090587

9. Gutierrez A, Norby FL, Maheshwari A, Rooney MR, Gottesman RF, Mosley TH, Lutsey PL, Oldenburg N, Soliman EZ, Alonso A, et al. Association of Abnormal P-Wave Indices With Dementia and Cognitive Decline Over 25 Years: ARIC-NCS (The Atherosclerosis Risk in Communities Neurocognitive Study). Journal of the American Heart Association. 2019;8:e014553. doi: 10.1161/jaha.119.014553

10. Karadag B, Ozyigit T, Ozben B, Kayaoglu S, Altuntas Y. Relationship between left atrial volume index and cognitive decline in elderly patients with sinus rhythm. Journal of Clinical Neuroscience. 2013;20:1074–1078. doi: 10.1016/j.jocn.2012.10.021

11. Martínez-González MA, Buil-Cosiales P, Corella D, Bulló M, Fitó M, Vioque J, Romaguera D, Martínez JA, Wärnberg J, López-Miranda J, et al. Cohort Profile: Design and methods of the PREDIMED-Plus randomized trial. International Journal of Epidemiology. 2018;48:387–388o. doi: 10.1093/ije/dyy225

12. Martínez-González MA, Buil-Cosiales P, Corella D, Bulló M, Fitó M, Vioque J, Romaguera D, Martínez JA, Wärnberg J, López-Miranda J, et al. Cohort Profile: Design and methods of the PREDIMED-Plus randomized trial. Int J Epidemiol. 2019;48:387–388o. doi: 10.1093/ije/dyy225

13. Upton J. Mini-Mental State Examination. In: Gellman MD, Turner JR, eds. Encyclopedia of Behavioral Medicine. New York, NY: Springer New York; 2013:1248–1249.

14. Folstein MF, Folstein SE, McHugh PR. “Mini-mental state”. A practical method for grading the cognitive state of patients for the clinician. Journal of Psychiatric Research. 1975;12:189–198. doi: 10.1016/0022-3956(75)90026-6

15. Mitchell AJ. A meta-analysis of the accuracy of the mini-mental state examination in the detection of dementia and mild cognitive impairment. Journal of Psychiatric Research. 2009;43:411–431.

16. Wambach D, Lamar M, Swenson R, Penney DL, Kaplan E, Libon DJ. Digit Span. In: Kreutzer JS, DeLuca J, Caplan B, eds. Encyclopedia of Clinical Neuropsychology. New York, NY: Springer New York; 2011:844–849.

17. Wechsler D. WAIS-III, escala de inteligencia de Wechsler para adultos-III : manual de aplicación y corrección. Madrid: TEA; 2001.

18. Patterson J. Controlled Oral Word Association Test. In: Kreutzer JS, DeLuca J, Caplan B, eds. Encyclopedia of Clinical Neuropsychology. New York, NY: Springer New York; 2011:703–706.

19. Peña-Casanova J, Quiñones-Ubeda S, Gramunt-Fombuena N, Quintana-Aparicio M, Aguilar M, Badenes D, Cerulla N, Molinuevo JL, Ruiz E, Robles A, et al. Spanish Multicenter Normative Studies (NEURONORMA Project): norms for verbal fluency tests. Archives of Clinical Neuropsychology. 2009;24:395–411. doi: 10.1093/arclin/acp042

20. Llinàs-Reglà J, Vilalta-Franch J, López-Pousa S, Calvó-Perxas L, Torrents Rodas D, Garre-Olmo J. The Trail Making Test. Assessment. 2017;24:183–196. doi: 10.1177/1073191115602552

21. Nagueh SF, Smiseth OA, Appleton CP, Byrd BF, 3rd, Dokainish H, Edvardsen T, Flachskampf FA, Gillebert TC, Klein AL, Lancellotti P, et al. Recommendations for the Evaluation of Left Ventricular Diastolic Function by Echocardiography: An Update from the American Society of Echocardiography and the European Association of Cardiovascular Imaging. Journal of the American Society of Echocardiography : official publication of the American Society of Echocardiography. 2016;29:277–314. doi: 10.1016/j.echo.2016.01.011

22. Thomas L, Hoy M, Byth K, Schiller NB. The left atrial function index: a rhythm independent marker of atrial function. Eur J Echocardiogr. 2008;9:356–362. doi: 10.1016/j.euje.2007.06.002

23. Machino-Ohtsuka T, Seo Y, Tada H, Ishizu T, Machino T, Yamasaki H, Igarashi M, Xu D, Sekiguchi Y, Aonuma K. Left atrial stiffness relates to left ventricular diastolic dysfunction and recurrence after pulmonary vein isolation for atrial fibrillation. J Cardiovasc Electrophysiol. 2011;22:999–1006. doi: 10.1111/j.1540-8167.2011.02049.x

24. Lip GY, Nieuwlaat R, Pisters R, Lane DA, Crijns HJ. Refining clinical risk stratification for predicting stroke and thromboembolism in atrial fibrillation using a novel risk factor-based approach: the euro heart survey on atrial fibrillation. Chest. 2010;137:263–272. doi: 10.1378/chest.09-1584

25. Sofi F, Macchi C, Abbate R, Gensini GF, Casini A. Mediterranean diet and health status: an updated meta-analysis and a proposal for a literature-based adherence score. Public Health Nutrition. 2014;17:2769–2782. doi: 10.1017/S1368980013003169

26. Elosua R, Marrugat J, Molina L, Pons S, Pujol E. Validation of the Minnesota Leisure Time Physical Activity Questionnaire in Spanish Men. American Journal of Epidemiology. 1994;139:1197–1209. doi: 10.1093/oxfordjournals.aje.a116966

27. Song R, Xu H, Dintica CS, Pan K-Y, Qi X, Buchman AS, Bennett DA, Xu W. Associations Between Cardiovascular Risk, Structural Brain Changes, and Cognitive Decline. Journal of the American College of Cardiology. 2020;75:2525–2534. doi: https://doi.org/10.1016/j.jacc.2020.03.053

28. Szegedi I, Szapáry L, Csécsei P, Csanádi Z, Csiba L. Potential Biological Markers of Atrial Fibrillation: A Chance to Prevent Cryptogenic Stroke. BioMed research international. 2017;2017:8153024–8153024. doi: 10.1155/2017/8153024

29. Russo C, Jin Z, Liu R, Iwata S, Tugcu A, Yoshita M, Homma S, Elkind MSV, Rundek T, Decarli C, et al. LA volumes and reservoir function are associated with subclinical cerebrovascular disease: the CABL (Cardiovascular Abnormalities and Brain Lesions) study. JACC Cardiovascular imaging. 2013;6:313–323. doi: 10.1016/j.jcmg.2012.10.019

30. Kresge HA, Khan OA, Wagener MA, Liu D, Terry JG, Nair S, Cambronero FE, Gifford KA, Osborn KE, Hohman TJ, et al. Subclinical Compromise in Cardiac Strain Relates to Lower Cognitive Performances in Older Adults. Journal of the American Heart Association. 2018;7:e007562. doi: 10.1161/JAHA.117.007562

31. Tsang TS, Abhayaratna WP, Barnes ME, Miyasaka Y, Gersh BJ, Bailey KR, Cha SS, Seward JB. Prediction of cardiovascular outcomes with left atrial size: is volume superior to area or diameter? J Am Coll Cardiol. 2006;47:1018–1023. doi: 10.1016/j.jacc.2005.08.077

32. Zhang MJ, Norby FL, Lutsey PL, Mosley TH, Cogswell RJ, Konety SH, Chao TF, Shah AM, Solomon SD, Alonso A, et al. Association of Left Atrial Enlargement and Atrial Fibrillation With Cognitive Function and Decline: The ARIC-NCS. Journal of the American Heart Association. 2019;8:e013197. doi: 10.1161/JAHA.119.013197

33. Shibuya-Tayoshi S, Sumitani S, Kikuchi K, Tanaka T, Tayoshi SY, Ueno S-I, Ohmori T. Activation of the prefrontal cortex during the Trail-Making Test detected with multichannel near-infrared spectroscopy. Psychiatry and Clinical Neurosciences. 2007;61:616–621. doi: https://doi.org/10.1111/j.1440-1819.2007.01727.x

34. Machino-Ohtsuka T, Seo Y, Tada H, Ishizu T, Machino T, Yamasaki H, Igarashi M, Xu D, Sekiguchi Y, Aonuma K. Left atrial stiffness relates to left ventricular diastolic dysfunction and recurrence after pulmonary vein isolation for atrial fibrillation. Journal of Cardiovascular Electrophysiology. 2011;22:999–1006. doi: 10.1111/j.1540-8167.2011.02049.x

